# Two-fold increase in the prevalence of autism in Israel between 2017 and 2021

**DOI:** 10.1101/2023.04.02.23287784

**Authors:** Ilan Dinstein, Shirley Solomon, Michael Zats, Ronit Shusel, Raphael Lottner, Bella Ben Gershon, Gal Meiri, Idan Menashe, Dorit Shmueli

## Abstract

The prevalence of autism spectrum disorder (ASD) has increased steadily in most high-income countries over the last decade. Estimates of ASD prevalence among 8-year-old children in the U.S. have increased from 1.47% in 2010 to 2.3% in 2018, and up to 3.5% among 3-17-year-old children in 2020. In Israel, estimates have been lower, with the prevalence of ASD in 8-year-old children increasing from 0.3% in 2009 to 0.64% in 2018. Here, we examined data from the entire population of Israel (∼3 million 1-17-year-old children) and quantified changes in their ASD prevalence between 2017 and 2021. We analyzed consecutive annual reports acquired from the Israeli National Insurance Institute (NII), which monitors all children with ASD in Israel who receive welfare services, and Clalit Health Services (CHS), the largest Health Maintenance Organization (HMO) in Israel that services ∼50% of the population. Both data sources revealed a nearly twofold increase in the ASD prevalence of 1–17-year-old children during this five-year period. Rates differed across age groups with 2-3-year-old (day-care) children exhibiting a ∼4.4-fold increase in prevalence from 0.27% to 1.19% and 4-6-year-old (pre-school) children exhibiting a ∼2.3-fold increase from 0.80% to 1.83%. These results demonstrate that ASD prevalence in Israel is increasing at unprecedented rates and shifting towards diagnosis at earlier ages. These findings highlight the challenge facing health and education service providers in meeting the needs of a rapidly growing ASD population that is being diagnosed at earlier ages.

## Introduction

Worldwide ASD prevalence in children under the age of 18 was recently estimated at 0.6-1%, on average (Salari et al., 2022; Zeidan et al., 2022). However, ASD prevalence estimates differ across age groups and vary widely across countries, regions, and ethnic populations. For example, in 2015 the ASD prevalence in children 7-9 years-old from Finland, Denmark, and Iceland was 0.77%, 1.26%, and 3.1%, respectively (Delobel-Ayoub et al., 2020a), while in 2016 the ASD prevalence in children 8 years-old from Colorado and New Jersey in the U.S. was 1.3% and 3.1%, respectively (Maenner et al., 2020). A variety of factors are likely to contribute to variable prevalence estimates across geographic locations including differences in social stigma, public awareness, availability and quality of health services, and accuracy of population monitoring techniques (Chiarotti & Venerosi, 2020; Leonard et al., 2010a). Additional variability is likely due to differences in population genetics and/or exposure to environmental factors (Gaugler et al., 2014; Rzhetsky et al., 2014).

While ASD prevalence may vary across countries for multiple reasons, a recent study has estimated that global prevalence has increased from ∼0.6% to ∼1% in the last decade (Zeidan et al., 2022). In the U.S. ASD prevalence has increased from 1.47% in 2010 to 2.3% in 2018 for 8-year-old children (Maenner et al., 2021) with an additional increase to 3.5% in 2020 for 3-17-year-old children (Li et al., 2022). In Canada prevalence has increased from 1.5% in 2015 to 2.2% in 2019 for 5–17-year-old children (Palmeter, O’Donnell, Lagace, Gheorghe, & Krupovich, 2019). Accurately quantifying ASD prevalence and its change over time is critical for planning policy and services. The importance of this is apparent in the establishment of longitudinal monitoring projects such as the Autism and Developmental Disabilities Monitoring (ADDM) Network by the Center for Disease Control (CDC) in the U.S. (Maenner et al., 2021).

In Israel, previous reports based on NII records have estimated that the national ASD prevalence among 8-year-old children was 0.12% in 2005, 0.3% in 2008, and 0.65% in 2015 (Gal, Abiri, Reichenberg, Gabis, & Gross, 2012; Segev, Weisskopf, Levine, Pinto, & Raz, 2019). This suggests a continuous increase in ASD prevalence with an overall 5.5-fold increase over ten years. An additional study using Maccabi Healthcare records, the second largest HMO in Israel (servicing ∼25% of the population), has reported higher rates of ASD prevalence among 8-year-old children with an almost three-fold increase from 0.46% in 2007 to 1.3% in 2018 (Davidovitch, Slobodin, Weisskopf, & Rotem, 2020). The difference between Maccabi and NII estimates may be due to different ethnic and socio-economic composition of the sub-populations insured by Maccabi HMO relative to the entire population of Israel. For example, several studies have reported that higher socio-economic abilities are associated with higher ASD prevalence in Israel (Davidovitch, Hemo, Manning-Courtney, & Fombonne, 2013; Raz, Weisskopf, Davidovitch, Pinto, & Levine, 2015; Segev et al., 2019).

The purpose of the current study was to quantify changes in ASD prevalence during the last 4 years and determine whether rates are continuing to increase or have stabilized as suggested by one previous study (Segev et al., 2019). Moreover, we wanted to determine whether prevalence changes differed across age groups and to specifically quantify prevalence increases in 2-3-year-old and 4-6-year-old children who are eligible for day-care and pre-school services, respectively. The Israeli healthcare and education systems offer comprehensive early intervention services to ASD children placed in either special or mainstream education settings (Ilan et al., 2021) and accurate estimates of ASD prevalence at these age groups are particularly important for planning the development of these services, given their importance for improving later outcomes (Gabbay-Dizdar et al., 2022; Zwaigenbaum et al., 2015). Accurate estimates are also important for properly budgeting rapid early diagnosis services and a wide range of additional ASD services given the large costs of ASD care (Buescher, Cidav, Knapp, & Mandell, 2014), with the average annual family expenditure in Israel estimated at over $8,000 (USD) in 2013 (Raz, Lerner-Geva, Leon, Chodick, & Gabis, 2013).

## Methods

### Study design and data sources

ASD prevalence was estimated based on data acquired from two independent sources: the National Insurance Institute (NII) of Israel and Clalit Healthcare Services (CHS) HMO. Israel has a universal, nationally funded healthcare system where the Ministry of Health is responsible for regulating health services that are supplied by four HMOs. Every Israeli resident is required to be registered with one of the four HMOs. CHS is the largest HMO, providing services to over half (52.4%) of the Israeli population (∼4.9 million residents in 2021). The NII is the national social security organization that implements the welfare policy of the Israeli government. Since 1981, the NII has provided financial support to families of children with ASD from the age of diagnosis and until the age of 18. The NII collects data on all Israeli residents (∼9.36 million residents in 2021), enabling the assessment of national ASD prevalence. The data analyzed in the current study comes from five annual reports generated by CHS and NII from their respective computerized records for the years 2017 to 2021.

### Case ascertainment by NII

To receive financial support from the NII, parents of children with ASD are required to submit a claim after their child receives the ASD diagnosis. Israeli regulations require that the ASD diagnosis be given by both a licensed psychologist and a physician (child psychiatrist or pediatric neurologist) according to DSM-5 criteria. The diagnosis must include physical, neurological, and developmental assessments with a detailed report of the child’s symptoms and functional, developmental, and cognitive status. All claims are reviewed by a professional committee at the NII to determine eligibility. As of January 2022, benefits included a monthly payment of NIS 3000 (US$965 in 2022) from the date of diagnosis until the age of 18. In addition, NII approval allows parents to receive tax deductions and access to early educational services (e.g., daycare special education). It is, therefore, assumed that the vast majority of ASD children are registered at the NII. Nevertheless, ASD related healthcare services at the HMOs may be accessed by individuals who have received the diagnosis but have not registered at the NII.

### Case ascertainment by CHS

To receive ASD related health services from CHS, a child must receive a formal diagnosis of ASD as described in the previous section. Diagnosed children are eligible for speech, occupational, behavioral, and psychological therapy as well as physiotherapy at HMO facilities or associated service providers. It is assumed that the vast majority of families with ASD children who are insured by CHS will report the ASD diagnosis to CHS in order to receive ASD related health services. However, ASD diagnoses can also be completed in private clinical settings without reporting the results to CHS. In such cases the families may apply for and receive NII support without any record of the ASD diagnosis at their HMO.

### Study Population

The study population included all 1–17-year-old children who were residents of Israel in each of the study years (2017-2021). Total resident numbers for each year were obtained from the Israeli Central Bureau of Statistics (CBS) which calculates population estimates based on census surveys (https://www.cbs.gov.il/en/Pages/search/yearly.aspx). The data includes population estimates by age, thereby allowing us to compute ASD prevalence for children at different ages. The total number of 1-17-year-old children in Israel (in thousands) was 3014.5 in 2017, 3071.5 in 2018, 3125.7 in 2019, 3239.3 in 2020, and 3360.2 in 2021.

### Data Analysis

We computed the annual prevalence of ASD from 2017 to 2021 in NII and CHS data separately. Prevalence estimates using NII data were calculated by dividing the number of 1–17-year-old children with an ASD diagnosis by the number of 1–17-year-old residents in Israel, separately in each calendar year. Prevalence estimates using CHS data were calculated by dividing the number of 1–17-year-old children with an ASD diagnosis in CHS records by the number of 1–17-year-old residents who were insured by CHS, separately in each calendar year. The number of Israeli residents insured by CHS was extracted from NII public data (https://www.btl.gov.il/Mediniyut/Situation/haveruth1/Pages/default.aspx). Equivalent analyses were performed for sub-groups of children binned by age or age groups corresponding to day-care (1-3 years), preschool (4-6 years), primary school (7-12 years), or middle/high school (13-17 years) educational settings. When assessing changes over time (e.g., annual increase rates) we excluded children under one year of age given the negligible number of ASD cases in this age group.

### Statistics

All statistical analyses were performed with the R statistical software (R Core Team, 2021). We performed a linear regression analysis with year and data source as predictors along with an additional interaction predictor for ‘year*source’ to assess whether the slope of prevalence changes differed across NII and CHS data. We performed a one-way ANOVA to test for differences in ASD prevalence across age groups with age, year of data, and source of data as main factors in the analysis. P-values <0.05 were considered significant. We also performed Z-tests of proportions to assess whether there were differences in the increase of ASD prevalence between NII and CHS from 2017 to 2021 for each of the 16 age groups. Results of these Z-tests were Bonferroni-corrected for 16 comparisons such that only P-values <0.003 were considered statistically significant.

## Results

According to NII data, the number of individuals with a formal diagnosis of ASD, 1-17-years-old, who were eligible for services increased from 14,914 in 2017 to 32,222 in 2021. This corresponds to nearly a 2-fold increase in ASD prevalence from 0.49% to 0.96% over the five-year period (Figure 1). Data from CHS suggested a slightly lower increase in prevalence from 0.48% in 2017 to 0.89% in 2021. A linear regression analysis demonstrated a highly significant increase in prevalence across years (β=0.0012; p<0.0001), as well as a significant difference in prevalence between NII and CHS (β=0.3629; p=0.0304) and a significant interaction between data source (NII and CHS data) and time/years (p = 0.0302), suggesting a steeper increase of ASD prevalence rates in the NII data compared to CHS data.

**Figure 1.**
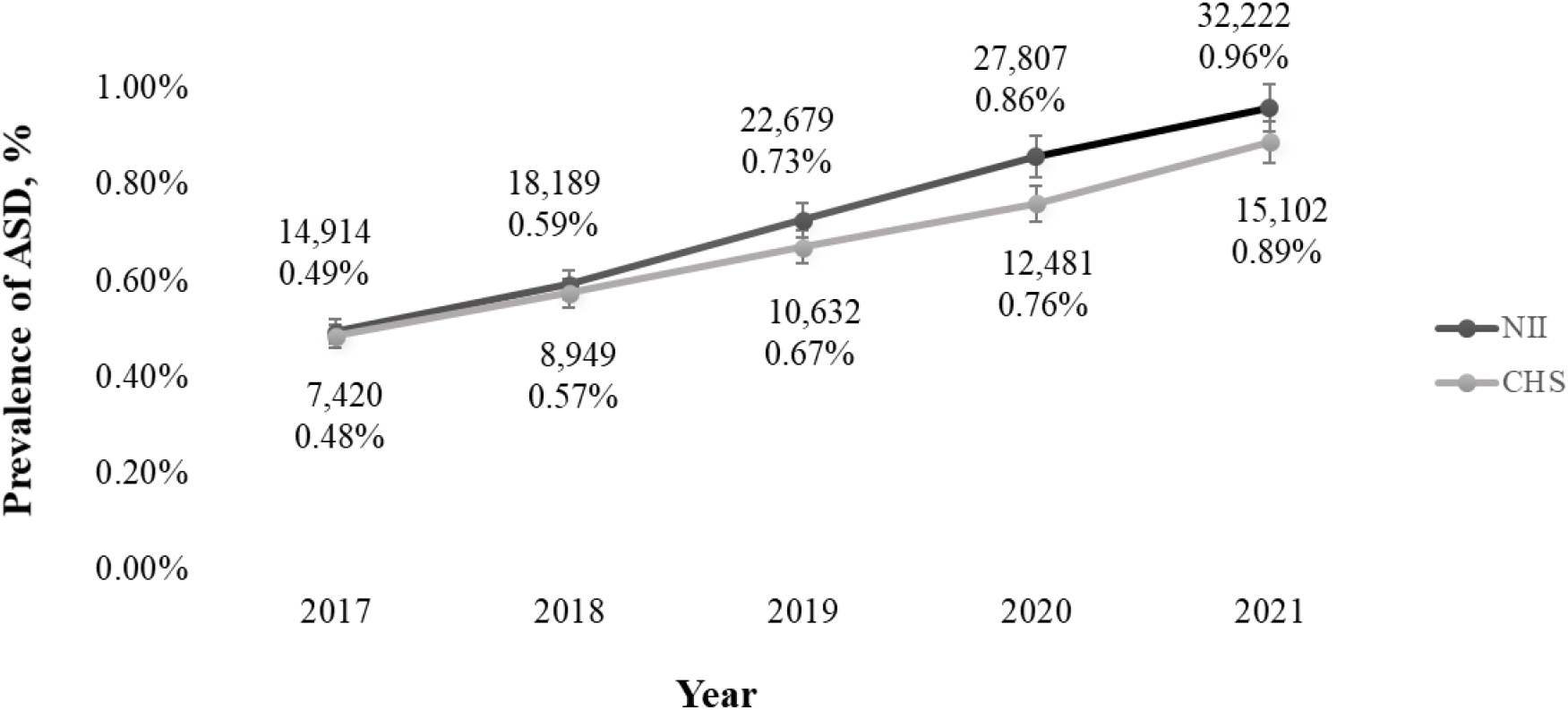
ASD prevalence rates in 1–17-year-old children as computed using NII (black) and CHS (gray) data between 2017 and 2021. Error bars represent the standard error.

Next, we compared ASD prevalence rates across age groups (Figure 2). This demonstrated that ASD prevalence was consistently highest in preschool children (4-6-years-old) regardless of the year (from 2017 to 2021). A one-way ANOVA revealed statistically significant differences in ASD prevalence (Figure 3, F(3,4)= 65.4; p= 0.0007) across four age groups: children in daycare (2-3-year-old), preschool (4-6-year-old), primary school (7-12 year old), and middle/high school (13-17 year old). Tukey’s test for multiple comparisons demonstrated that ASD prevalence was significantly higher in the preschool group relative to the daycare (p = 0.0101, 95% C.I. = 0.0021, 0.0048), primary school (p = 0.0009, 95% C.I. = 0.0013, 0.0057), and middle/high school (p = 0.0477, 95% C.I. = 0.0032, 0.0044) groups. ASD prevalence was also significantly higher in the primary school group relative to day care (p = 0.0007, 95% C.I. = 0.0048, 0.0091) and middle/high school (p = 0.0015, 95% C.I. = 0.0035, 0.0079) groups. There was also a significant effect for data source (F(1,4) = 12.22; p=0.0396), with NII data yielding higher estimates of ASD prevalence than CHS data.

**Figure 2.**
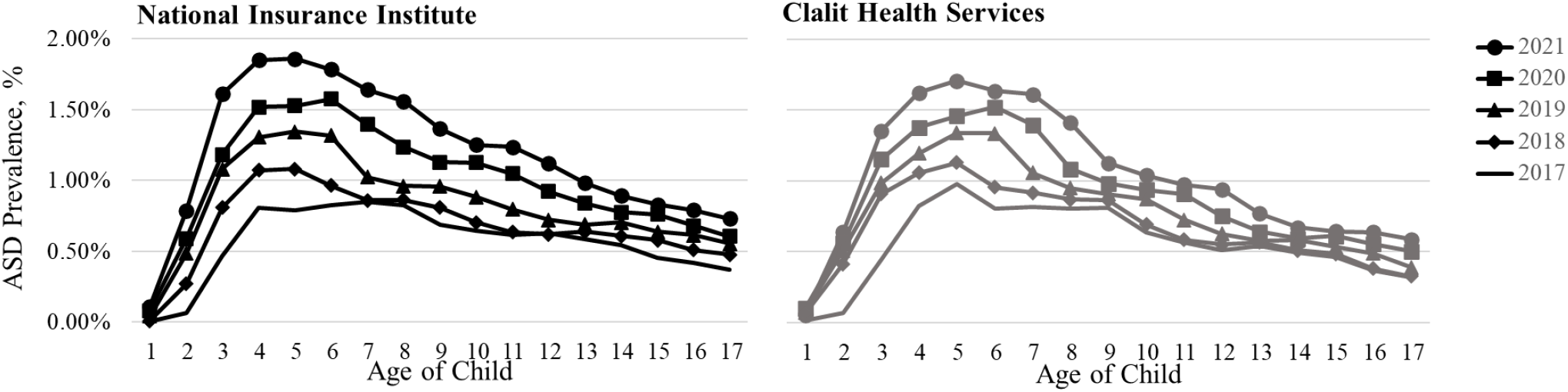
Prevalence of ASD by age as computed from NII (left, black) and CHS (right, gray) data for 2017 (solid line), 2018 (diamonds), 2019 (triangles), 2020 (squares) and 2021 (circles).

**Figure 3.**
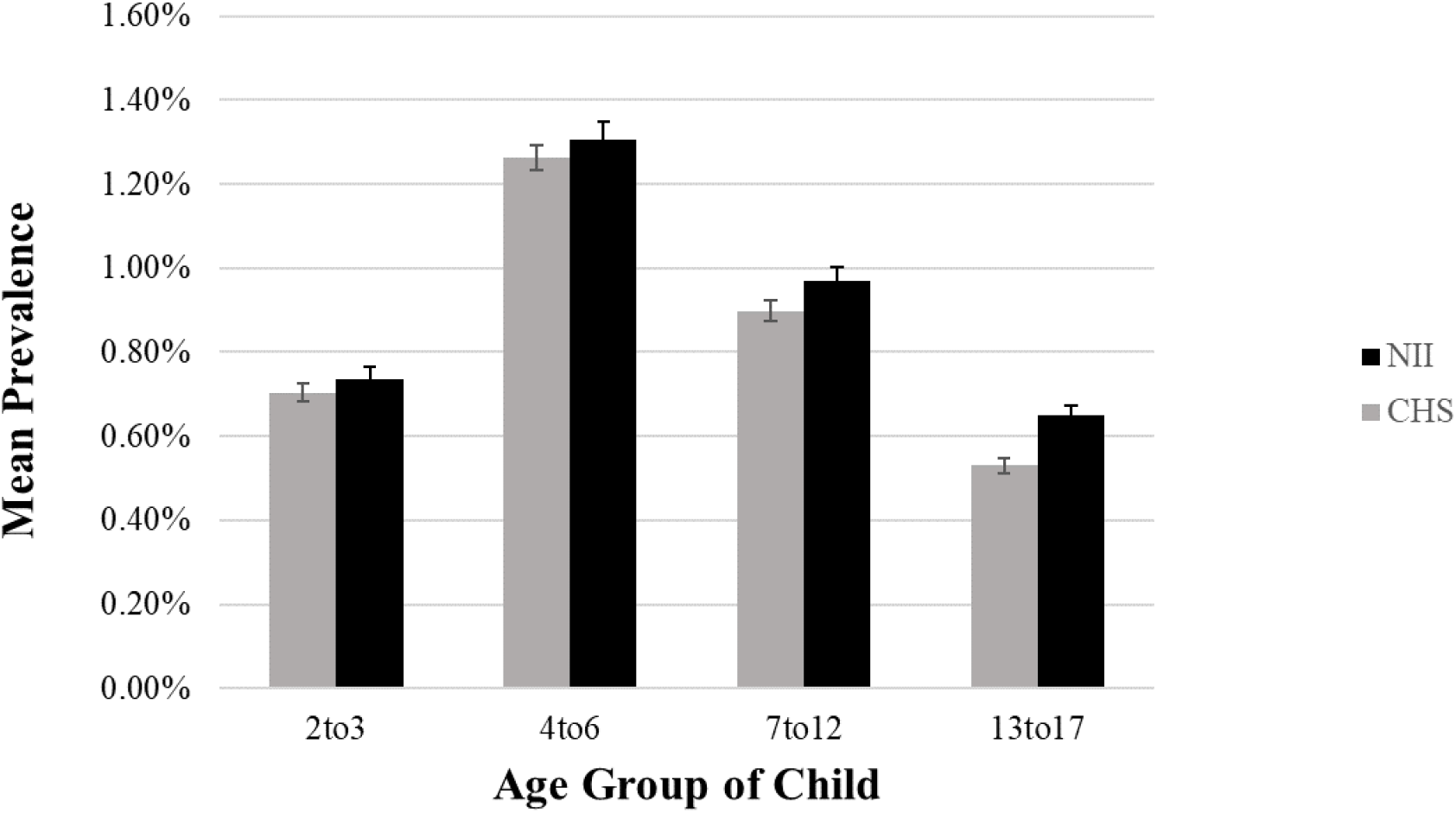
Mean ASD prevalence in daycare (2-3-year-old), preschool (4-6-year-old), primary school (7-12-year-old), and middle/high school (13-17) children between 2017 and 2021. Error bars: standard error of the mean. Black: NII data. Gray: CHS data.

ASD prevalence increased annually in all age groups from 2017 to 2021, but not at the same rate (Figure 4). The largest increases were apparent in children 3 to 7 years old, with mean annual increases of 0.16-0.23% and 0.15-0.18% reported in NII and CHS data, respectively. In contrast, 10-year-old children exhibited annual increases that were approximately half as large and annual increases were even smaller for older children. While annual increases in prevalence were overall larger according to NII data (M[SD] = 0.13%[0.0009] vs. 0.09%[0.0103]), there were no significant differences across NII and CHS data by age after Bonferroni correction for multiple comparisons (p>0.003).

**Figure 4.**
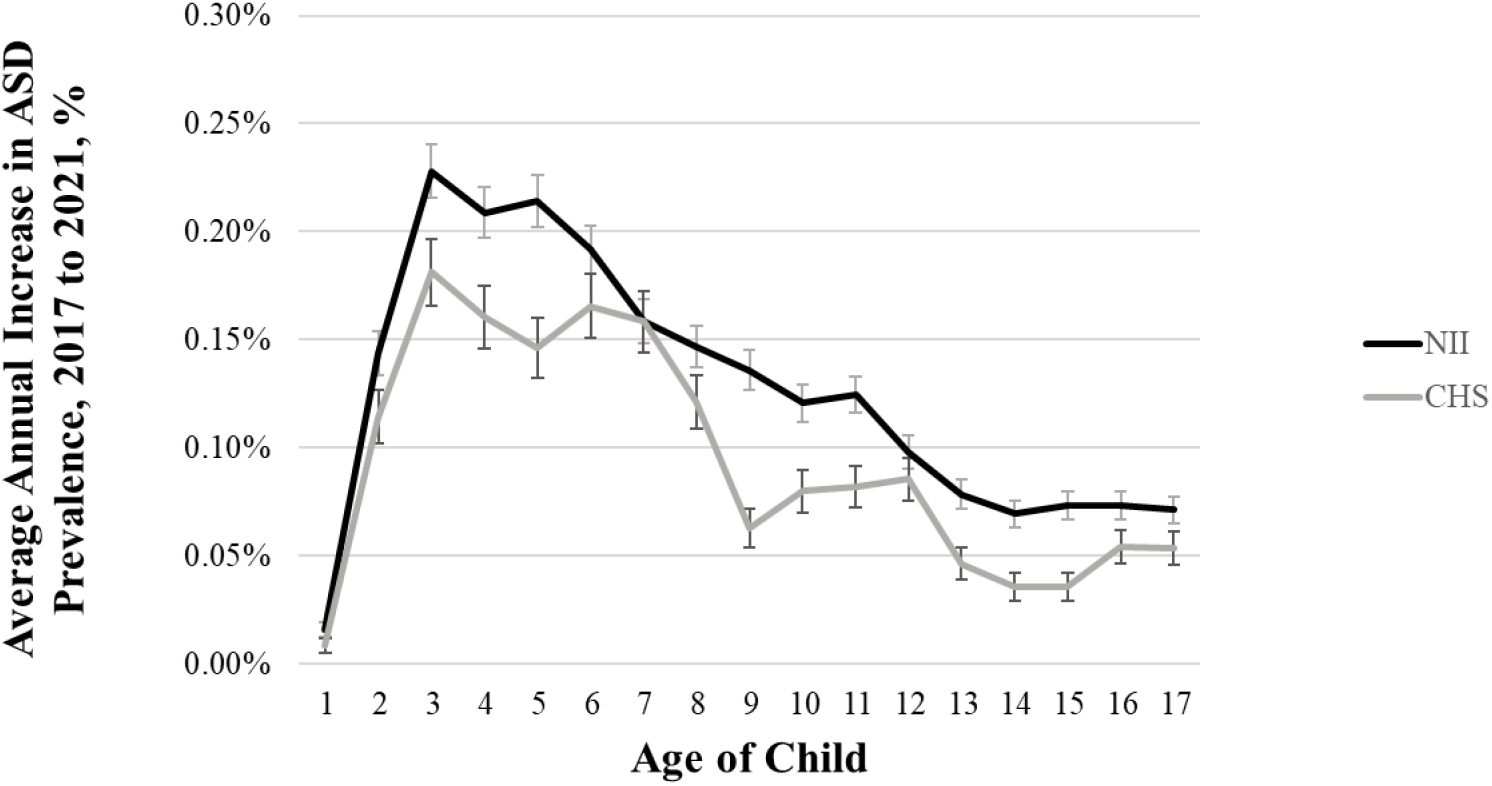
Mean annual increase in ASD prevalence per age from 2017 to 2021 as deduced from NII (black) and CHS (gray) data. Error bars: standard error of the mean.

We also calculated the total fold increase in ASD prevalence from 2017 to 2021 per age (Figure 5). The greatest fold increase was found at age 2 with a 12 and 9.4-fold increase in NII and CHS data, respectively. This was followed by 3-year-old children who exhibited a 3.5 and 3-fold increase in NII and CHS data, respectively. In contrast, the total fold increase from 2017 to 2021 remained relatively stable in older children ages 4 to 17 with an average of 1.96 (IQR = 0.04) in NII data and 1.70 (IQR = 0.06) in CHS data. This demonstrates the shift in ASD diagnosis to earlier ages (i.e., 2–3-year-olds).

**Figure 5.**
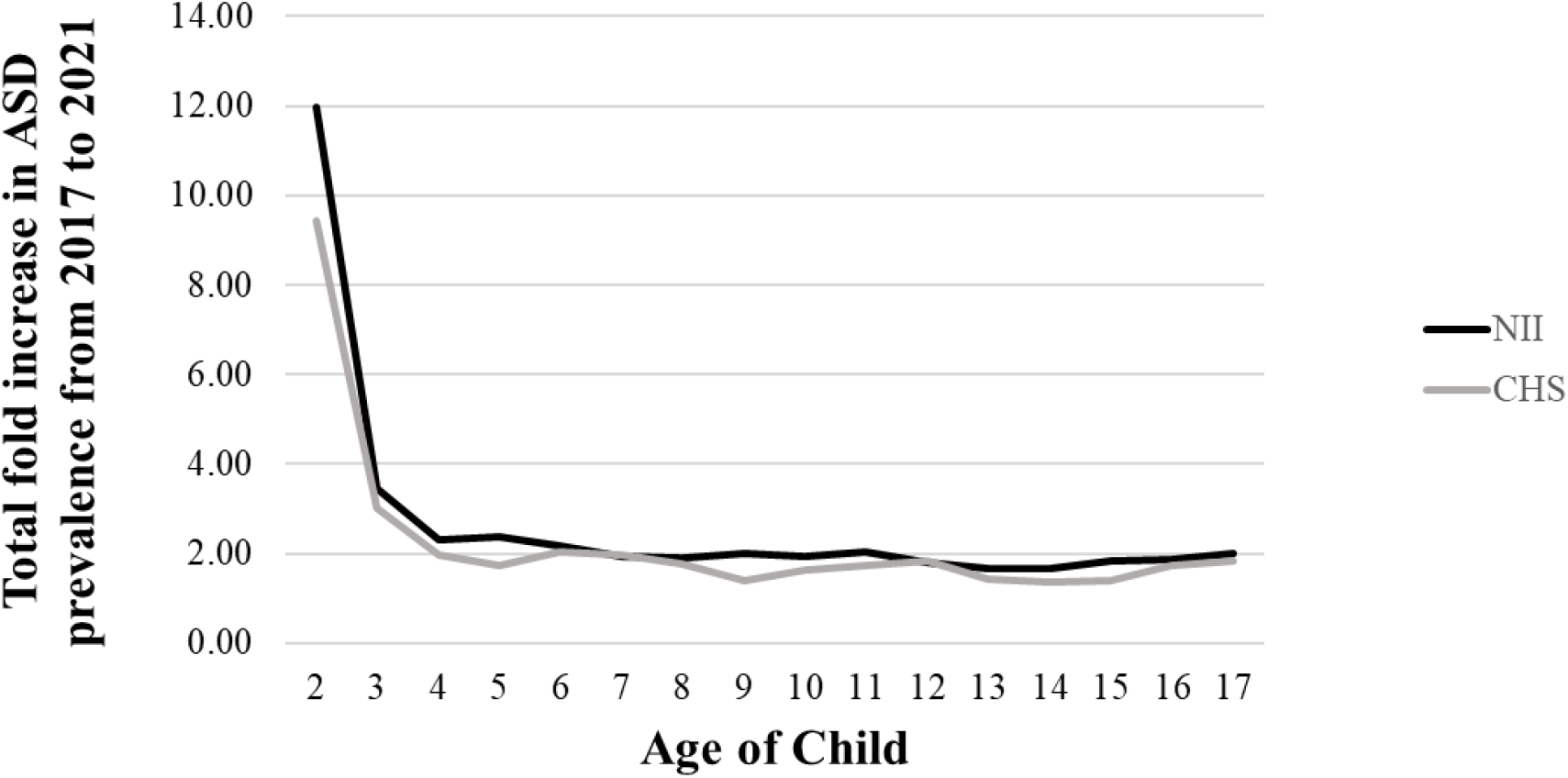
Total fold increase in ASD prevalence from 2017 to 2021 per age group in NII (black) and CHS (gray) data.

In a final analysis we re-examined the changes in ASD prevalence separately for daycare, preschool, primary school, and middle/high school age groups (Figure 6). We performed equivalent linear regression analyses for each of the age groups, which revealed the following. For daycare children there was a significant increase in the prevalence across years (β= 0.0022; p= 0.0002) with no significant differences in prevalence between NII and CHS (β= 1.0696; p= 0.2044) and no interaction between data source (NII and CHS data) and time/years (p = 0.2042). For preschool children there was a significant increase in the prevalence across years (β= 0.0025; p<0.0001) along with a significant difference in prevalence between NII and CHS (β= 1.150; p= 0.0005) and a significant interaction between data source (NII and CHS data) and time/years (p= 0.0005), such that prevalence rates increased significantly more in NII relative to CHS data. For primary school children there was a significant increase in prevalence across years (β= 0.0017; p= 0.0001) with no significant differences in prevalence between NII and CHS (β= 0.9160; p= 0.1538) and no interaction between data source (NII and CHS data) and time/years (p= 0.1535). Finally, for middle/high school children there was a significant increase in prevalence across years (β= 0.0009; p<0.0001) along with a significant difference in prevalence between NII and CHS (β= 0.6505; p= 0.0052) and a significant interaction between data source (NII and CHS data) and time/years (p= 0.0051), indicating that prevalence rates increased significantly more in NII data.

**Figure 6.**
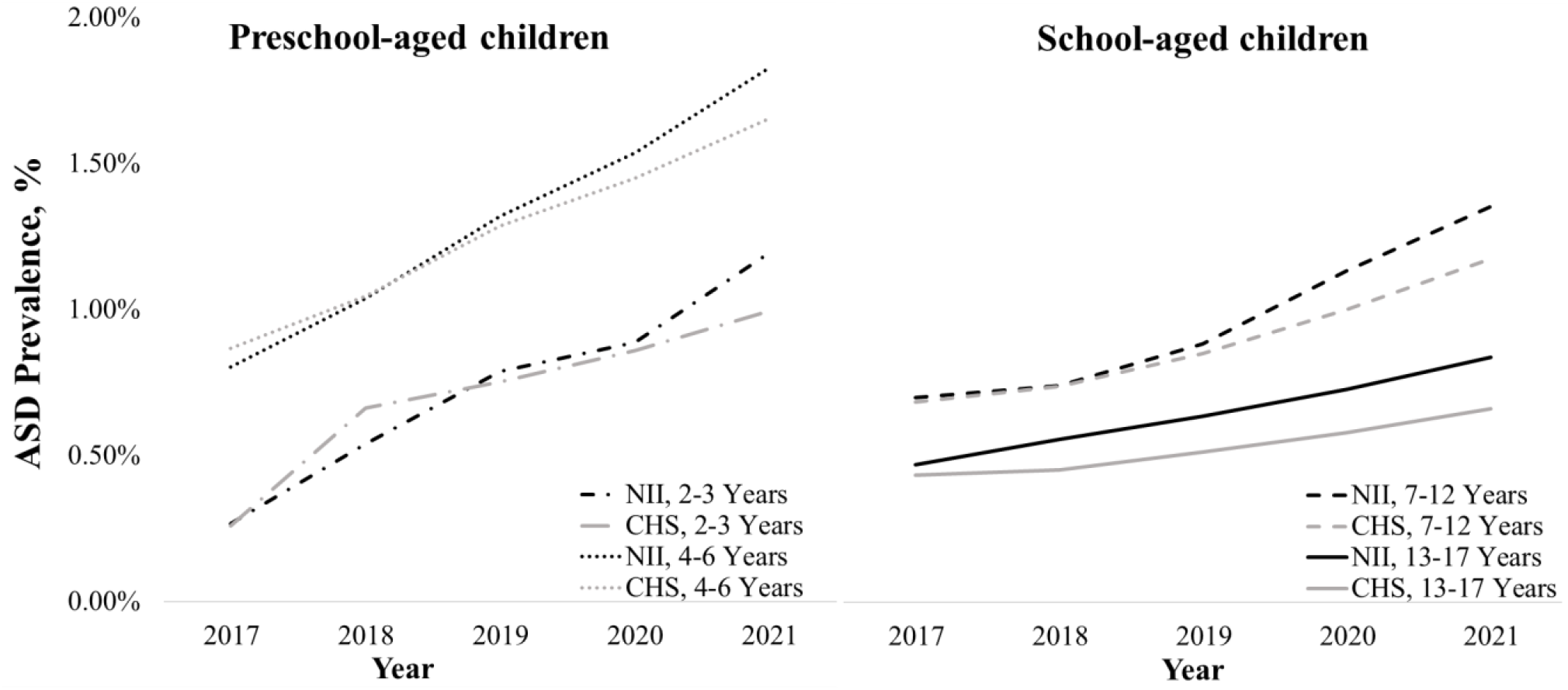
Changes in ASD prevalence from 2017 to 2021 in each of four age groups: daycare (2-3 years old), preschool (4-6 years old), primary school (7-12 years old), and middle/high school (13-17 years old). NII (black) and CHS (gray) data are presented in separate lines.

## Discussion

Our results demonstrate that ASD prevalence continued to grow rapidly in Israel between 2017 and 2021. While overall ASD prevalence in children 1-17-years-old increased by a factor of two (Figure 1), ASD prevalence in children 2-3-years-old increased by a factor of 4.4. This demonstrates a rapid shift towards early ASD diagnosis in Israel such that in 2021 the ASD prevalence was 1% among 2-3-year-old children and almost 2% among 4-5-year-old children (Figure 5). The high percentage of diagnosed children at such young ages creates an opportunity for early intervention that is important for improving later outcomes (Gabbay-Dizdar et al., 2022; Zwaigenbaum et al., 2015). However, capitalizing on this opportunity requires a corresponding rapid increase in the availability of early intervention services that are managed by the Israel healthcare and education systems.

Since children rarely loose an ASD diagnosis (Wiggins et al., 2012), these findings suggest that ASD prevalence in older children and adolescents in the Israeli population will reach at least 2% within a decade as the ASD children described in the current study grow older and are joined by those diagnosed at older ages. These results, therefore, highlight the need to expand the healthcare, educational, and social services necessary to support this population as it ages.

### ASD prevalence in Israel relative to other countries

Previous estimates of ASD prevalence in Israel have been considerably lower than those reported in most comparable high-income countries around the world. While ASD prevalence among 8-year-old children in the U.S. was already 1.47% in 2010 (Maenner et al., 2021), ASD prevalence for this age group in Israel was only 0.65% in 2015 (Segev et al., 2019). Our results demonstrate that it has increased to 1.5% in 2021 (Figure 2). This suggests that ASD had been previously under-diagnosed in Israel and that the Israeli healthcare system is closing the gap towards ASD prevalence rates described in other countries.

Since ASD prevalence is affected by a complex combination of social, political, ethnic, financial, genetic, and environmental factors (Leonard et al., 2010b), it is difficult to predict whether ASD prevalence in Israel will continue to increase at a similar rapid rate over the next 4-5 years or start to stabilize. Note that in some countries such as Iceland (Delobel-Ayoub et al., 2020b) and U.S. states such as California and New Jersey (Maenner et al., 2021), ASD prevalence among 8-year-old children has already exceeded 3%. Hence, ASD prevalence is likely to continue and grow in Israel if one assumes that there is a similar percentage of children with ASD across different geographic locations and that population genetics and environmental differences have only minor impact on ASD prevalence.

### Generalizability of ASD prevalence estimates across data sources

Our results demonstrate the importance of using complete national data for accurately estimating ASD prevalence and its change over time. The rate of increase in ASD prevalence differed significantly across estimates from NII and CHS data (Figure 1). While the general trend was equivalent, ASD prevalence increased more modestly in CHS data relative to NII data. This suggests two possible interpretations, which are not mutually exclusive. The first is that more families with newly diagnosed children were registering with NII than with CHS systems, perhaps due to a larger urgency of receiving benefits and access to early education services (NII jurisdiction) rather than ASD-related healthcare services (CHS jurisdiction). The second is that the national increase in ASD prevalence may be driven more strongly by diagnosis rates of individuals insured by other HMOs rather than those of CHS. Indeed, a recent study with Maccabi HMO data (the second largest HMO in Israel, servicing ∼25% of the population), reported an ASD prevalence rate of 1.3% among 8-year-old children already in 2018 (Davidovitch et al., 2020). This ASD prevalence was ∼0.5% higher than that reported by CHS for the equivalent year. We speculate that these differences may be due to the ethnic and socio-economic characteristics of those insured by the different HMOs, given that CHS insures more of the marginalized ethnic populations in the geographic periphery of Israel (Davidovitch et al., 2013; Raz et al., 2015; Segev et al., 2019).

### Growth of health, education, and social services in corresponding years

There is limited publicly available data about the corresponding growth of ASD related services in Israel, making it difficult to quantify the response of public systems to the growth in ASD prevalence. Nevertheless, according to NII data, the number of families receiving supplemental income due to ASD diagnoses in Israel has doubled between 2017 and 2021 (Figure 1). Moreover, according to Israeli law, children with ASD (unlike children with ID or other developmental disorders) are eligible for intensive early intervention programs at the ages of 1-7-years-old. According to health ministry data (personal communication, unpublished), the utilized budget of these programs has doubled from 255 million NIS in 2017 to 510 million NIS in 2021 (287 in 2018, 400 in 2019, and 447 in 2020, in millions NIS). While ASD prevalence at these ages has more than doubled during this period (Figure 4), it is remarkable that the availability of early intervention programs is growing at such a rapid pace in Israel. This is due to the intervention programs that are run by the Ministry of Health via non-government organization such as ALUT and the association for children at risk. According to data from the ministry of education (personal communication, unpublished), the number of special education kindergartens for ASD children 3-6-years-old has grown from 625 in 2020 to 723 in 2021, indicating an annual increase of ∼15.6%, which is also remarkable, but still lower than the annual rate of prevalence increase (∼21%, Figure 3) for this age group.

### Limitations

The current study had several limitations. First, we did not have information regarding the sex or geographic composition of ASD cases, which limited our ability to identify prevalence differences across males and females or distinct geographic areas of Israel. Second, we did not have retrospective data from before 2017, which limited our ability to calculate cumulative incidence rates by birth cohort as reported by previous studies in Israel (Raz et al., 2015; Segev et al., 2019). However, given that ASD diagnoses are rarely lost (Wiggins et al., 2012), ASD prevalence rates are likely to be equivalent to cumulative incidence rates such that, for example, ASD prevalence of 4-year-olds in 2017 will be equivalent to the ASD cumulative incidence rate of those born in 2013. Third, we did not have equivalent data from Maccabi, Meuhedet, and Leumit HMOs that service ∼50% of Israeli residents. This limited our ability to assess HMO specific differences and ASD prevalence changes.

## Conclusions

ASD prevalence in 1-17-year-old children has doubled in Israel between 2017 and 2021 and quadrupled in 2-3-year-old children. This rapid growth has considerable healthcare, educational, and social ramifications, which are currently met only partially. These findings highlight the necessity of a coordinated effort to further expand support services for this population at multiple ages. Improving public availability of NII, HMO, Health Ministry, and Education Ministry data will greatly facilitate the assessment of ASD prevalence in Israel and its growth over time to better plan future services for this population.

## Data Availability

All data produced in the present work are contained in the manuscript

